# Wastewater Surveillance for Monkeypox Virus in Nine California Communities

**DOI:** 10.1101/2022.09.06.22279312

**Authors:** Marlene K. Wolfe, Alexander T. Yu, Dorothea Duong, Madhura S. Rane, Bridgette Hughes, Vikram Chan-Herur, Marisa Donnelly, Shua Chai, Bradley J. White, Duc J. Vugia, Alexandria B. Boehm

## Abstract

**Background:** Wastewater represents a composite biological sample from the entire contributing population. People infected with monkeypox virus (MPXV)^1^ may excrete viral DNA into wastewater via multiple ways such as in feces, urine, skin lesions, and/or saliva. We describe results from rapid establishment of wastewater surveillance in selected regions in California within a month of the first reported case of monkeypox in the United States.

**Methods:** PCR assays targeting genomic DNA from MPXV were deployed in an ongoing wastewater surveillance program in California. MPXV DNA concentrations were measured daily in settled solids samples from nine wastewater plants. Results over a four-week period were validated across different MPXV assays, compared using influent and solids samples, and correlated using non-parametric methods (Kendall’s tau) with the number of monkeypox cases reported from each sewershed.

**Results:** MPXV DNA was detected at all nine sites between June 19 and August 1, 2022; 5 of 9 sites detected MPXV DNA prior to or within a day of the first case identified in the source sewershed. At the four sites with >10 positive detections, we observed a positive, statistically significant correlation (p <0.001) between MPXV DNA in wastewater solids and incidence rate of reported cases.

**Conclusions:** Our findings suggest wastewater can be used to effectively detect the introduction of MPXV and monitor its circulation in the community to inform public health and clinical response. This flexible wastewater surveillance infrastructure may be rapidly leveraged to monitor other pathogens of public health importance that are shed into wastewater.

## Introduction

Monkeypox virus (MPXV), of the *Orthopoxvirus* genus in the family *Poxviridae*, is endemic in Western and Central Africa where infection has been linked to transmission from infected animals or humans. Sporadic cases and outbreaks linked to travel or imported animals have been recognized in non-endemic countries since the first identification of human disease in 1970^1^. In May 2022, cases of monkeypox infection without association to endemic areas were reported in multiple European countries^1^. Within weeks, cases were identified in the United States and a dozen countries around the world. By late July, the World Health Organization declared monkeypox a public health emergency of international concern^2^. Unlike previous outbreaks, this global outbreak is driven by human-to-human transmission and cases without association to each other have been identified, suggesting additional undetected community transmission^3,4^.

Since recognition of the global outbreak, there has been a rapid scale-up of public health response, including substantial increases in testing and efforts to educate clinicians and the public to mitigate spread^5^. However, surveillance is dependent on practical access to and utilization of testing, limited by awareness of a disease that is novel to the general public and most clinicians, stigma of a disease that has to date primarily been diagnosed in gay, bisexual, and other men who have sex with men, and the potential for asymptomatic cases^6,7^. Alternative public health surveillance approaches independent of individual testing, such as wastewater surveillance, provide an attractive means to detect and track emerging monkeypox transmission and provide situational awareness for public health agencies and clinicians.

Use of wastewater surveillance to monitor trends in infectious diseases has grown rapidly. Wastewater represents a composite biological sample combining inputs from community members connected to a sewer network and many pathogens are shed in ways that reliably end up in wastewater, including in urine, feces, oral and nasal secretions, and sloughing of skin. Wastewater surveillance has been reliably used by public health agencies throughout the coronavirus disease 2019 (COVID-19) pandemic to monitor for SARS-CoV-2, the causative virus of COVID-19. Concentrations of SARS-CoV-2 viral RNA are strongly correlated with COVID-19 case incidence^8–10^, and recent studies show this is also the case for other respiratory viruses such as Influenza A and respiratory syncytial virus^11,12^.

Recent small observational studies of monkeypox infection have confirmed the presence of viral DNA for some individuals in saliva, feces, urine, semen, and/or skin lesions^13,14^. Although concentrations of viral DNA in specimens were not reported, Cq values (a measurement of how many cycles are needed to detect a signal) from quantitative polymerase chain reaction (PCR) measurements were low, suggesting high concentrations of MPXV DNA even 16 days after symptom onset^13^. These data are consistent with reports of MPXV shedding from previous outbreaks in humans^15^ and experimental studies in animals^16–19^. Related viruses including smallpox have been shown to be excreted in urine^20^. Together, this evidence suggests that MPXV DNA is likely to appear in wastewater. Based on a systematic review of the literature^21^, no study to date has documented the persistence of orthopoxviruses in wastewater. However, Vaccinia virus has been reported to persist for days in raw freshwater and marine waters^21^, suggesting orthopoxviruses may persist as wastewater transits from homes to nearby wastewater treatment plants.

In a collaborative effort between researchers running a wastewater surveillance program and the California Department of Public Health (CDPH), we rapidly adapted and deployed an assay for surveillance of MPXV DNA in multiple sewersheds in California. We describe results from this work, including the establishment of monitoring within a month of the first reported case of monkeypox in the US and an exploration of the relationship between MPXV DNA concentrations in wastewater and cases in the community.

## Methods

### MPXV Molecular Assay

To detect MPXV DNA in wastewater, we used assays developed by the United States Centers for Disease Control and Prevention (CDC)^22^. The G2R_G assay, which targets a region of the OPG002 gene common to all MPXV sequences, was used on all samples. A subset of samples (described further below) was also assayed for MPXV DNA using the G2R_WA assay which targets a separate region of the OPG002 gene that is specific to Clade II. The choice of assays was confirmed by alignment with sequences from the 2022 outbreak.

### Wastewater samples for MPXV surveillance

Each day between June 19, 2022 and August 1, 2022, settled solids were collected from nine publicly owned treatment works (POTWs) in California in the Greater San Francisco Bay and Sacramento areas (Table 1, S1). The POTWs treat wastewater from between 66,622 and 1,480,00 people; details of the POTWs and specific sample collection processes are provided in Wolfe et al.^8^ Samples were couriered to a laboratory the same day they were collected and processed immediately, unless otherwise specified (Table S2), with results available within 24 hours of sample collection. A total of 407 solids samples were included in this study.

**Table 1.**
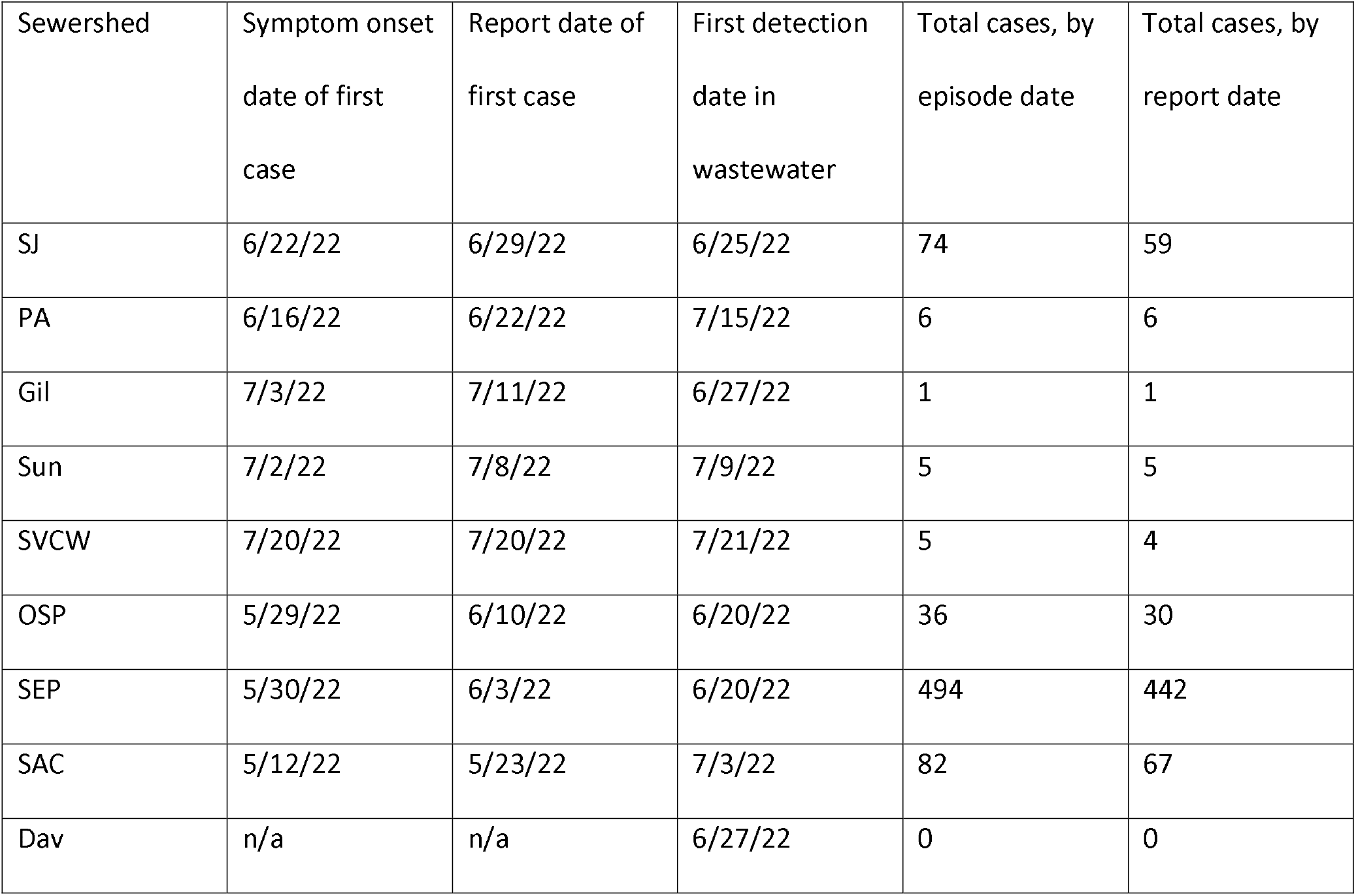
First cases of monkeypox and first detection of monkeypox virus (MPXV) DNA in wastewater by sewershed, California. Sewersheds serviced by publicly owned treatment works (POTWs): SJ (San Jose), PA (Palo Alto), Gil (Gilroy), Sun (Sunnyvale), SVCW (Silicon Valley Clean Water), OSP (Oceanside, San Francisco), SEP (Southeast, San Francisco), Sac (Sacramento), Dav (Davis). Symptom onset date of the first case in each sewershed was available and thus reported. Total cases using episode date (symptom onset date, or if not available, the earliest of laboratory result or case record creation date) and report date are noted separately for each sewershed within the study period (June 19 to August 1, 2022).

| Sewershed | Symptom onset<br>date of first<br>case | Report date of<br>first case | First detection<br>date in<br>wastewater | Total cases, by<br>episode date | Total cases, by<br>report date |
| --- | --- | --- | --- | --- | --- |
| SJ | 6/22/22 | 6/29/22 | 6/25/22 | 74 | 59 |
| PA | 6/16/22 | 6/22/22 | 7/15/22 | 6 | 6 |
| Gil | 7/3/22 | 7/11/22 | 6/27/22 | 1 | 1 |
| Sun | 7/2/22 | 7/8/22 | 7/9/22 | 5 | 5 |
| SVCW | 7/20/22 | 7/20/22 | 7/21/22 | 5 | 4 |
| OSP | 5/29/22 | 6/10/22 | 6/20/22 | 36 | 30 |
| SEP | 5/30/22 | 6/3/22 | 6/20/22 | 494 | 442 |
| SAC | 5/12/22 | 5/23/22 | 7/3/22 | 82 | 67 |
| Dav | n/a | n/a | 6/27/22 | 0 | 0 |

At the laboratory, nucleic acids were extracted and purified from the solids using previously published methods^8,23–25^ (see SI for additional details). Nucleic acids were used undiluted as template in digital droplet PCR wells; 10 replicate wells were run per sample and results from the wells were merged to determine the concentration of the G2R_G target. PCR cycling conditions and further details of digital droplet PCR (ddPCR) are provided in the SI. We also measured pepper mild mottle virus (PMMoV) RNA gene concentrations; as well as recovery of spiked-in bovine coronavirus (BCoV) RNA following methods outlined elsewhere^23–25^. Additionally, negative and positive extraction controls, and negative and positive PCR controls were included in each plate. Gene fragments were used as controls for the G2R_G assay; other controls are described elsewhere^8^. Nucleic acids were not stored prior to analysis, with the exception of some from a POTW serving the part of San Francisco (SEP) which were stored at -80°C less than two weeks and subjected to a freeze thaw prior to G2R_G analysis (Table S2).

A subset of samples with the highest concentrations of G2R_G detection from the two POTWs with the most frequent G2R_G (OSP and SEP, Table S2) detections were processed for the second MPXV genomic target, G2R_WA, using the same replication and QA/QC as described for G2R_G. A gene fragment was used as the G2R_WA positive control. PCR cycling conditions are provided in the SI.

### Liquid influent samples

Samples of 24-h composited influent were collected in sterile containers on ∼7 consecutive days (Table S2) from two POTWs (OSP and SEP) that had the highest rates of detection of the G2R_G target to compare the concentrations obtained using liquid wastewater to those obtained using settled solids. Methodological details are in the SI. In brief, viral targets were concentrated from wastewater influent using Nanotrap particles (Ceres Nanosciences, Manassas, VA) before nucleic acids were extracted, purified, and used as template in digital droplet PCR to measure concentrations of G2R_G and G2R_WA targets, as well as PMMoV and BCoV recovery following the same protocol as described for solids.

### Case Data

Counts of incident cases of monkeypox, defined in this study as people with lesion swabs PCR positive for orthopoxvirus or MPXVs, were recorded as a function of episode date (symptom onset date, or if not available, the earliest of laboratory result or case record creation date) and report date (laboratory result date or, if not available, case record create date). Cases were aggregated within sewersheds based on georeferenced home addresses, delineated using POTW-specific GIS shape files. To obtain daily incidence rates of monkeypox, a 7-day rolling average of incident cases per 100,000 population was calculated using the estimated population served in each sewershed (Table S1). To explore incidence of cases of monkeypox at the time MPXV DNA was first detected in wastewater at each sewershed, we compared incidence during periods with no MPXV detection (any 7-day period without detection in wastewater) with the period of first detection (first 7-day period with at least 2 detections in wastewater). Weekly incidence of cases of monkeypox was estimated for each of these periods (all cases with an episode date within 7-day periods of no detection of MPXV DNA, or all cases within the 7-day period leading up to the date of first detection of MPXV DNA, divided by total sewershed population). Since the goal was to describe upper limits of incidence of monkeypox cases in each sewershed when MPXV DNA was not detected in wastewater, for sewersheds with multiple 7-day periods without detection of MPXV DNA in wastewater, the single highest weekly incidence of cases of monkeypox was used.

### Statistical analysis

We assessed the association between the moving 5-day trimmed (highest and lowest of the 5 values excluded) average of MPXV DNA concentrations in wastewater solids and 7-day moving averages of monkeypox daily incidence using Kendall’s tau for the subset of sites for which both variables had more than 10 values (out of ∼44) greater than 0 to ensure both variables had sufficient variance. Tests were repeated using the MPXV DNA concentrations in wastewater normalized by PMMoV, and for both the episode date and report date associated with cases. Non-parametric methods were chosen as data were not normally distributed (Shapiro-Wilk test). We also assessed the relationship between MPXV DNA concentrations in measurements taken from liquid and solids samples and the relationship between results from the G2R_G and G2R_WA assays on the same sample using Kendall’s tau. A paired Wilcoxon Sign Rank test was then used to assess the difference in results between liquids and solids, and between the G2R_G and G2R_WA assays used on the same samples. All statistical analysis was performed in RStudio (version 2021.09.2).

ab

This activity was reviewed by CDC and was conducted consistent with applicable federal law and CDC policy.§

## Results

MPXV DNA was detected in wastewater samples across all sites (9/9) monitored during the study period from June 19 to August 1, 2022 (Table 1, Fig 1, Fig 2, Fig S4). Concentrations of MPXV DNA (G2R_G target) ranged from non-detect to 24,113 copies/g dry weight of wastewater solids (more about lowest detectable concentration in SI). Positive and negative controls were all positive and negative respectively, BCoV recoveries were higher than 10%, and PMMoV concentrations were within an expected range for each POTW. This indicates assays performed efficiently and demonstrated no evidence of contamination. A subset of samples was run with a second assay (G2R_WA, specific to Clade II of MPXV) and no significant difference between measurements of G2R_WA and G2R_G were observed (details in SI). A comparison of results from liquid and solids samples showed a significant association (Kendall’s tau = 0.52, n = 28, p = 0.00012), with significantly higher concentration in solids (about 10^3^ higher) on a per mass basis (Wilcoxon signed rank test, n = 28, p<0.001) (Fig 3). Further details are in the SI.

**Figure 1.**
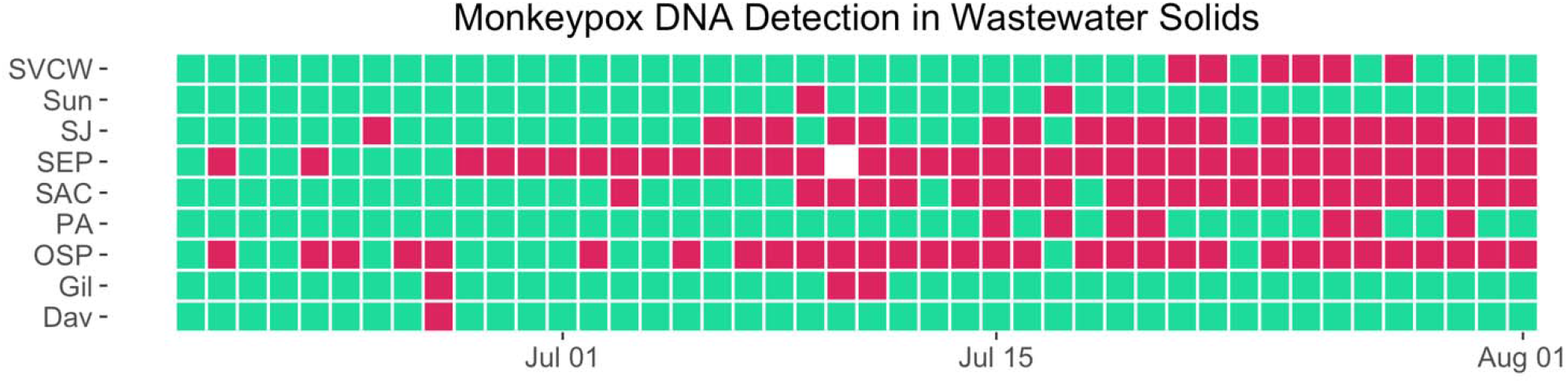
Monkeypox DNA detection in wastewater solids by sewershed. Heatmap of positive and negative results from all publicly owned treatment works (POTWs): SJ (San Jose), PA (Palo Alto), Gil (Gilroy), Sun (Sunnyvale), SVCW (Silicon Valley Clean Water), OSP (Oceanside, San Francisco), SEP (Southeast, San Francisco), Sac (Sacramento), Dav (Davis). Red indicates a positive detection, green indicates a negative detection. White indicates no sample collected.

**Figure 2.**
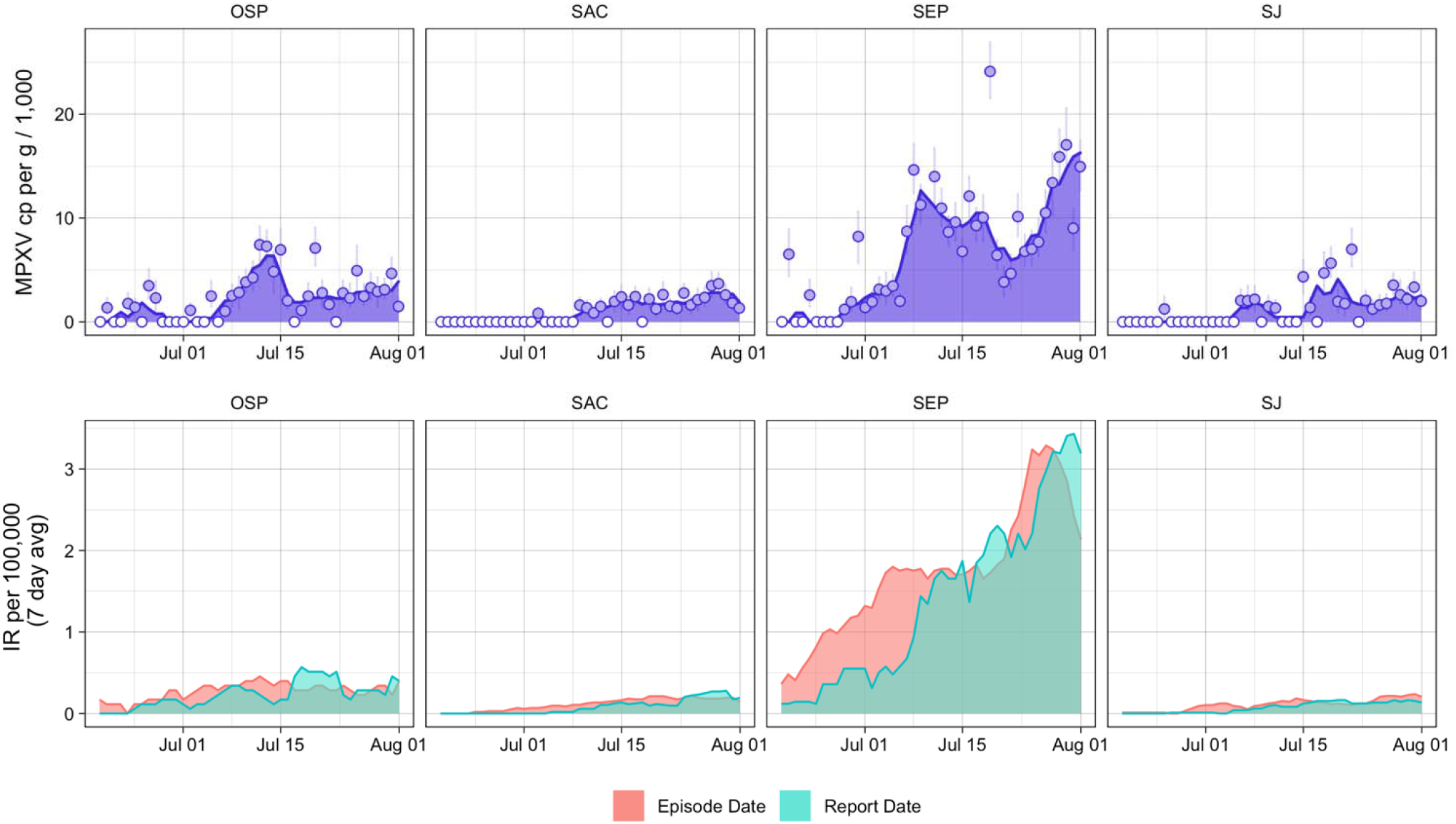
Wastewater monkeypox virus (MPXV) concentrations and incidence of monkeypox cases by sewershed. Top row: Time series of wastewater concentrations (concentration of MPXV DNA normalized by concentration of PMMoV RNA) at select publicly owned treatment works (POTWs) with >10 positive detections during the study time period: SEP (Southeast, San Francisco), OSP (Oceanside, San Francisco), Sac (Sacramento), and SJ (San Jose). The area under the curve represents the 5 day trimmed average of MPXV DNA cp/g over PMMoV cp/g in wastewater. Points represent daily values; open circles indicate non-detects. Error bars represent standard deviations and include Poisson error and variability among the 10 replicates (68% confidence intervals reported by the instrument software as “total error”). Bottom row: Daily incidence rate (IR) or monkeypox cases, averaged over 7-days, using episode date (red) and report date (green).

**Figure 3.**
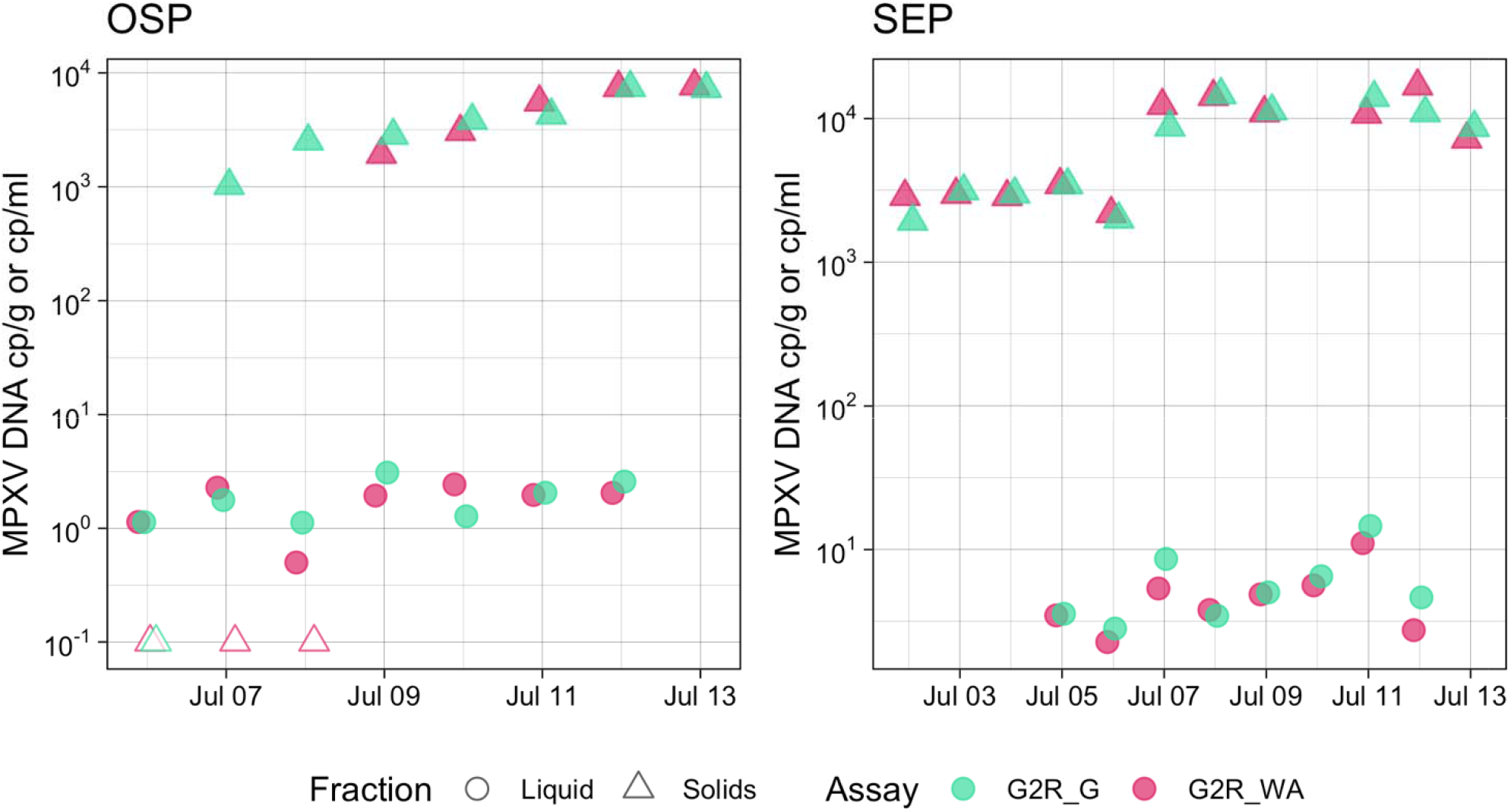
Concentrations of monkeypox virus (MPXV) in wastewater solids and liquid influent by assay. Concentrations of G2R_G and G2R_WA in wastewater solids and liquid influent at OSP (Oceanside, San Francisco) and SEP (Southeast, San Francisco) POTWs. Units are per gram for measurements using wastewater solids, and per mL for measurements from liquid wastewater.

The first clinical monkeypox case in any of the study areas was reported in the Sacramento sewershed on May 23, 2022; wastewater monitoring for MPXV began on June 19 (Table 1). The first wastewater samples to test positive were June 20 samples from each of the two facilities serving San Francisco (OSP, SEP). Daily samples from these sites were sporadically positive for the following 1-2 weeks, after which samples were consistently positive in increased concentrations (Fig 1, Fig 2, Fig S4). The next sites to test positive were SJ (June 25), and SAC (July 3). These sites also had a pattern of sporadic positives followed by increasing rates of positive samples and concentrations. Samples from SUN, PA, Gil, Dav, and SVCW also tested positive; SVCW and PA had some increase in rates of positive samples.

During the study period, the total number of cases in each sewershed ranged from 0 (Dav) to 494 (SEP). In SJ, Gil, and Dav, a first positive detection in wastewater preceded the report of a first case of monkeypox. During periods with no detection in wastewater, the highest weekly incidence recorded at each sewershed was an average of 0.68 cases (median 0.65, range 0-1.28) per 100,000. The average weekly incidence when a sewershed first had at least 2 detections in 7 days was 1.14 (median 0.95, range 0.89-2.01) per 100,000.

At the four sites with >10 positive samples and days with cases, there was a significant, positive association between the 5-day trimmed average concentration of viral DNA in wastewater and the 7-day average monkeypox incidence rate in the corresponding sewershed both when cases were compared using episode date (Kendall’s tau = 0.59, p <0.001, n = 176) and report date (Kendall’s tau = 0.66, p <0.001, n = 130). Results were similar when wastewater data were normalized by PMMoV and with raw, daily concentrations from wastewater (Table S4).

## Discussion

Surveillance for infectious diseases is a core function to inform public health, clinical, and general public understanding of risk and strategies for prevention of disease and is especially important for emerging infectious diseases. In the current global monkeypox outbreak, localized detection of disease introduction and circulation informs public health and clinical response, allows appropriate allocation of scarce testing, therapeutic and vaccine resources, and is important for risk messaging to the public. Traditional case surveillance for monkeypox is dependent on confirmatory PCR diagnostic testing of cases. While such testing is important for clinical decision making and is the backbone of disease surveillance, limitations include variable disease recognition, variable care-seeking due to disease severity and stigma, testing availability, and clinician awareness. Complementary strategies that can overcome these limitations and provide rapid population level awareness, like wastewater surveillance, can be vital for public health response.

We found that an established wastewater surveillance infrastructure could be rapidly leveraged to detect and monitor MPXV DNA to inform public health response. MPXV DNA was consistently detectable in wastewater from sewersheds with confirmed cases, even in locations with few identified cases. In some places, detections in wastewater preceded identification of the first cases in the community. We also found that the level of viral DNA in wastewater correlated with monkeypox incidence rate, suggesting that wastewater surveillance is a viable methodology to monitor trends in monkeypox disease activity. These findings suggest that wastewater surveillance can be adopted where feasible as an adjunct public health tool in the current global outbreak to monitor monkeypox disease activity, including in areas without known cases.

It is not yet possible to translate the concentration of MPXV DNA in wastewater to a predicted number of cases in the sewershed. One important reason is the unclear understanding of the true case incidence in each sewershed. However, comparisons between the weekly case incidence at the time of first consistent detections in wastewater (at least two detections within a 7-day period), and the weekly incidence during 7-day periods of no detection suggest that the beginning of consistent detections in wastewater correspond to a weekly incidence of at least 0.89-2.01 cases per 100,000. This is a likely underestimate, with true cases under-reported due to the reasons discussed above. One sewershed (Dav) had one positive wastewater sample but no reported cases; this likely reflects unidentified case(s) in residents or visitors.

There was a higher association between wastewater concentration and case incidence by report date, as compared to episode date (primarily reflecting symptom onset date). This suggests both that wastewater surveillance results appear timely relative to when cases are known to public health, as well as a lag between symptom onset and detection of MPXV DNA in wastewater, potentially reflecting delayed viral shedding into wastewater. Further research on MPXV shedding is necessary to describe how viral shedding may affect lag and vary by disease severity, and to improve estimates of the number of cases based on a wastewater concentration.

Wastewater samples regularly collected for routine public health monitoring can be tested for new targets with minimal change in processes. Our findings suggest wastewater can be used to effectively monitor for the introduction of MPXV and track its circulation for public health, clinical, and public awareness. The rapid adaptation of wastewater surveillance infrastructure to effectively monitor a non-enteric, non-respiratory virus such as MPXV, is promising for the future use of this tool for emerging infectious diseases of public health concern. Increased and sustained investment is needed to build wastewater surveillance infrastructure and for it to be ready to adapt rapidly to meet new and emerging public health needs.

## Data Availability

Wastewater data are publicly available in the Stanford Digital Repository (https://purl.stanford.edu/gz983hf3741)

## Acknowledgements

This work is supported by a gift from the CDC Foundation. Numerous people contributed to sample collection, including Srividhya Ramamoorthy (Sac), Michael Cook (Sac), Ursula Bigler (Sac), James Noss (Sac), Lisa C. Thompson (Sac), Payal Sarkar (SJ), Ryan Batjiaka (OSP and SEP), Melanie Wong (SEP), Lily Chan (Ocean), the Oceanside and Southeast plant operations personnel, Karin North (PA), Armando Guizar (PA), Saeid Vaziry (Gil), Chris Vasquez (Gil), Alo Kauravlla (Sun), Maria Gawat (SVCW), Tiffany Ishaya (SVCW), Eric Hansen (SVCW), and Jeromy Miller (Dav). We acknowledge Robert Snyder at California Department of Public Health and Caroline Sheikhzadeh from Emory University for their assistance. This study was performed on the ancestral and unceded lands of the Muwekma Ohlone people. We pay our respects to them and their Elders, past and present, and are grateful for the opportunity to live and work here.

Dorothea Duong, Bridgette Hughes, Vikram Chan-Herur, and Bradley White are employees of Verily Life Sciences.

## Disclaimer

The findings and conclusions in this report are those of the authors and do not necessarily represent the official positions of the California Department of Public Health or the California Health and Human Services Agency.

## Disclaimer

The findings and conclusions in this report are those of the authors and do not necessarily represent the official positions of the Centers for Disease Control and Prevention.

§ See e.g., 45 C.F.R. part 46, 21 C.F.R. part 56; 42 U.S.C. §241(d); 5 U.S.C. §552a; 44 U.S.C. §3501 et seq.

