## Supplementary Information for "Wastewater Surveillance for Monkeypox Virus in Nine California Communities"

**Brief solids processing methods.** In brief, dewatered solids were suspended in a buffer (75 mg / ml) and homogenized, and then 10 replicate aliquots of the buffer were subjected to nucleic acid extraction and purification, followed by inhibitor removal using commercial kits (Chemagic Viral DNA/RNA 300 Kit H96 for Chemagic 360 (PerkinElmer, Waltham, MA) and Zymo OneStep-96 PCR Inhibitor Removal kit (Zymo Research, Irvine, CA)). These methods have been published previously^1^.

**Influent processing methods.** Samples were couriered to the laboratory on the same day they were collected and stored at 4°C for between 1 and 7 d before being processed together in one batch. For each sample, 10 replicate aliquots were processed using an affinity-based capture method with magnetic hydrogel Nanotrap Particles with Enhancement Reagent 1 (Ceres Nanosciences, Manassas, VA) on 10mL of sample to concentrate viral particles using a KingFisher Flex system. RNA was then extracted from the each concentrated aliquot using the MagMAX Viral/Pathogen Nucleic Acid Isolation Kit (Applied Biosystems, Waltham, MA) on the KingFisher Flex platform to obtain purified nucleic acids which were then process through a Zymo OneStep-96 PCR Inhibitor Removal kit (Zymo Research, Irvine, CA)). Nucleic acids were used undiluted as template in digital droplet PCR to measure concentrations of G2R_G and G2R_WA targets, as well as PMMoV and BCoV recovery following the same protocol as described for the solids.

**G2R_G and G2R_WA ddPCR detailed methods.** dd-PCR was performed on 20 µl samples from a 22 µl reaction volume, prepared using 5.5 µl template, mixed with 5.5 µl of One-Step RT-ddPCR Advanced Kit for Probes (Bio-Rad 1863021), 2.2 µl of 200 U/µl Reverse Transcriptase, 1.1 µl of 300 mM DTT and primers and probes at a final concentration of 900 nM and 250 nM respectively. Primer and probes for assays were purchased from Integrated DNA Technologies (IDT, San Diego, CA) (Table S3). We used one step RT-ddPCR mastermix so that we could multiplex the samples with genomic RNA (gRNA) targets of other viruses as part of our regional wastewater monitoring program. The G2R_G assay was multiplexed with an assay targeting gRNA of human metapneumovirus and a mutation in SARS-CoV-2 Omicron BA.5 (HV69-70) (results for other assays not provided herein). The G2R_WA assay was multiplexed with assays targeting gRNA of human rhinovirus and influenza B virus (data from these RNA viruses is not included herein). We assayed four different concentrations of G2R_G standards and four nucleic-acid extracts from wastewater samples for G2R_G using ddPCR mastermix and one-step RT-ddPCR mastermix in conjunction with a RT step during thermocycling and found results did not differ (Figure S1).

Droplets were generated using the AutoDG Automated Droplet Generator (Bio-Rad, Hercules, CA). PCR was performed using Mastercycler Pro (Eppendforf, Enfield, CT) with with the following cycling conditions: reverse transcription at 50°C for 60 minutes, enzyme activation at 95°C for 5 minutes, 40 cycles of denaturation at 95°C for 30 seconds and annealing and extension at 59°C for 30 seconds, enzyme deactivation at 98°C for 10 minutes then an indefinite hold at 4°C. The ramp rate for temperature changes were set to 2°C/second and the final hold at 4°C was performed for a minimum of 30 minutes to allow the droplets to stabilize. Droplets were analyzed using the QX200 Droplet Reader (Bio-Rad). A well had to have over 10,000 droplets for inclusion in the analysis. All liquid transfers were performed using the Agilent Bravo (Agilent Technologies, Santa Clara, CA).

Thresholding was done using QuantaSoft™ Analysis Pro Software (Bio-Rad, version 1.0.596). In order for a sample to be recorded as positive, it had to have at least 3 positive droplets. Each wastewater sample was run in 10 replicate wells, and each 96-well PCR plate of wastewater samples included PCR positive controls for each target assayed on the plate in 1 well, and PCR NTCs in two wells. PCR positive controls consisted of gene fragments.

Results from replicate wells were merged for analysis. For the wastewater solid samples, three positive droplets across 10 merged wells corresponds to a concentration between ~500-1000 cp/g and this represents the lowest detectable concentration; the range in values is a result of the range in the equivalent mass of dry solids added to the wells. For the wastewater influent samples, three positive droplets across 10 merged wells corresponds to a concentration between ~1 cp/ml (the lowest detectable concentration).

Concentrations of RNA targets were converted to concentrations per dry weight of solids in units of copies/g dry weight or copies / ml for influent using dimensional analysis. The dry weight of the dewatered solids was determined by drying in an oven^2^. The total error is reported as standard deviations and includes the errors associated with the Poisson distribution and the variability among the 10 replicates.

**Supplementary Results**

Results from the G2R_G assay and the G2R_WA assay were significantly associated (Kendall’s tau = 0.88, n = 34, p < 0.001) and there was no significant difference between the two measurements (Wilcoxon signed rank test, n = 34, p = 0.16). Wastewater data are publicly available in the Stanford Digital Repository ([https://purl.stanford.edu/gz983hf3741](https://urldefense.com/v3/__https:/purl.stanford.edu/gz983hf3741__;!!AvL6XA!zOnlBi9MEJ1ydZnONp7zcJW2ch6Pr70ZEHUmlGUL6prjGRwAYe8y4qVxS0mm0OCc3oAQx7ko-RodfCOfUP2kXP6p$))

Our methods focus on the use of wastewater solids, as previous research indicates that many viruses are highly concentrated in solids in wastewater. We compared concentrations of MPXV DNA in paired liquid and solids samples from the two San Francisco sites over a period of one week. There was a significant association between the results from liquids and solids (Kendall’s tau = 0.52, n = 28, p = 0.00012). Concentrations of MPX DNA in solids by either assay were significantly higher than those in liquids on a per mass basis (Wilcoxon signed rank test, n = 28, p<0.001), with about a 10^3^ higher concentration of viral DNA observed per g or mL in solids.

The association between wastewater results and cases were also similar when wastewater was normalized by PMMoV, and when daily data was used for wastewater without smoothing (Table S4).

Table S1. Some POTW characteristics. See Wolfe et al.^1^ or [wbe.stanford.edu](http://wbe.stanford.edu) for more information.

| Plant names and abbreviations | County locations in California, USA | Population served |
| --- | --- | --- |
| San Jose (SJ) | Santa Clara County | 1,458,017 |
| Palo Alto (PA) | Santa Clara County | 213,968 |
| Gilroy (Gil) | Santa Clara County | 110,338 |
| Sunnyvale (Sun) | Santa Clara County | 169,000 |
| Silicon Valley Clean Water (SVCW) | San Mateo County | 220,000 |
| Oceanside (OSP) | San Francisco County | 250,000 |
| Southeast (SEP) | San Francisco County | 650,000 |
| Sacramento (SAC) | Sacramento County | 1,480,000 |
| Davis (Dav) | Davis County | 66,622 |

Table S2. Details of sample collection and storage for subset of samples from OSP (Oceanside, San Francisco) and SEP (Southeast, San Francisco) that were not immediately processed. In most cases, storage was required to run special comparison studies (solids versus influent, or assay GR2_G versus assay GR2_WA). In the case of SEP, some samples (14 nucleic acid extracts and 9 solids samples) used to generate the daily time series had to be stored due to lab and operational constraints. Note that all other sample used to generate the POTW time series (407-43= 364) were processed immediately upon collected. Dates are in month/day/year format. Samples were stored between 1 and 12 days, depending on sample) and analyzed on Sample Analysis Date listed. Based on previous work, storage of samples at the temperatures and times indicated is not expected to impact measurements ^3–5^

| Samples | Experiment | Storage Temp | Dates Collected (number of samples during period) | Sample Analysis Date |
| --- | --- | --- | --- | --- |
| Influent collected at OSP | Influent vs solids | 4°C | 7/6/22-7/12/22 (7) | 7/14/22 |
| Influent collected at SEP | Influent vs solids | 4°C | 7/5/22 - 7/12/22 (8) | 7/14/22 |
| SEP solids analyzed for G2R_WA | GR2_G vs GR2_WA | 4°C | 7/2/22 - 7/9/22, 7/11/22 - 7/13/22 (11) | 7/14/22 |
| OSP solids analyzed for G2R_WA | GR2_G vs GR2_WA | 4°C | 7/6/22 - 7/13/22 (8) | 7/14/22 |
| SEP solids nucleic acids run for G2R_G | Generation of daily MPXV time series | -80°C | 6/18/22 - 6/29/22 (12) and  7/2/22 - 7/4/22 (2) | 6/30/22  and  7/5/22 |
| SEP solids samples run for G2R_G | Generation of daily MPXV time series | 4°C | 7/5/22 - 7/11/22 (7)  and  7/12/22 – 7/18/22 (7)  And  7/19/22-7/25/22 (7)  And  7/26/22 – 8/1/22 (8) | 7/12/22  and  7/19/22  And  7/25/22  and  8/1/22 |

Table S3. Primer and probe sequences from Li et al^6^. FAM, 6-fluorescein amidite; HEX, hexachloro-fluorescein; ZEN, a proprietary internal quencher from IDT; 3IABkFQ, 3' Iowa Black FQ.

| Target | Primer/Probe | Sequence |
| --- | --- | --- |
| G2R_G | Forward | GGAAAATGTAAAGACAACGAATACAG |
|  | Reverse | GCTATCACATAATCTGGAAGCGTA |
|  | Probe | AAGCCGTAATCTATGTTGTCTATCGTGTCC (56-FAM/ZEN/3IABkFQ or 5HEX/ZEN/3IABkFQ) |
| G2R_WA | Forward | CACACCGTCTCTTCCACAGA |
|  | Reverse | GATACAGGTTAATTTCCACATCG |
|  | Probe | AACCCGTCGTAACCAGCAATACATTT  (56-FAM/ZEN/3IABkFQ or 5HEX/ZEN/3IABkFQ) |

Table S4. Results of Kendall’s tau test for the association between MPXV DNA in wastewater and reported monkeypox cases. Case data is utilized both by episode date and report date, and wastewater data is utilized as MPXV cp/g and MPXV normalized by PMMoV, and with and without 5-day trimmed smoothing.

| Wastewater Variable | Case Variable | Kendall’s Tau | p-value |
| --- | --- | --- | --- |
| 5-day trimmed avg MPXV cp/g | 7-day avg episode date | 0.57 | <0.001 |
| 5-day trimmed avg MPXV cp/g | 7-day avg report date | 0.68 | <0.001 |
| 5-day trimmed avg MPXV/PMMoV | 7-day avg episode date | 0.63 | <0.001 |
| 5-day trimmed avg MPXV/PMMoV | 7-day avg report date | 0.72 | <0.001 |
| Daily cp/g | 7-day avg episode date | 0.52 | <0.001 |
| Daily cp/g | 7-day avg report date | 0.56 | <0.001 |
| Daily MPXV/PMMoV | 7-day avg episode date | 0.52 | <0.001 |
| Daily MPXV/PMMoV | 7-day avg report date | 0.61 | <0.001 |

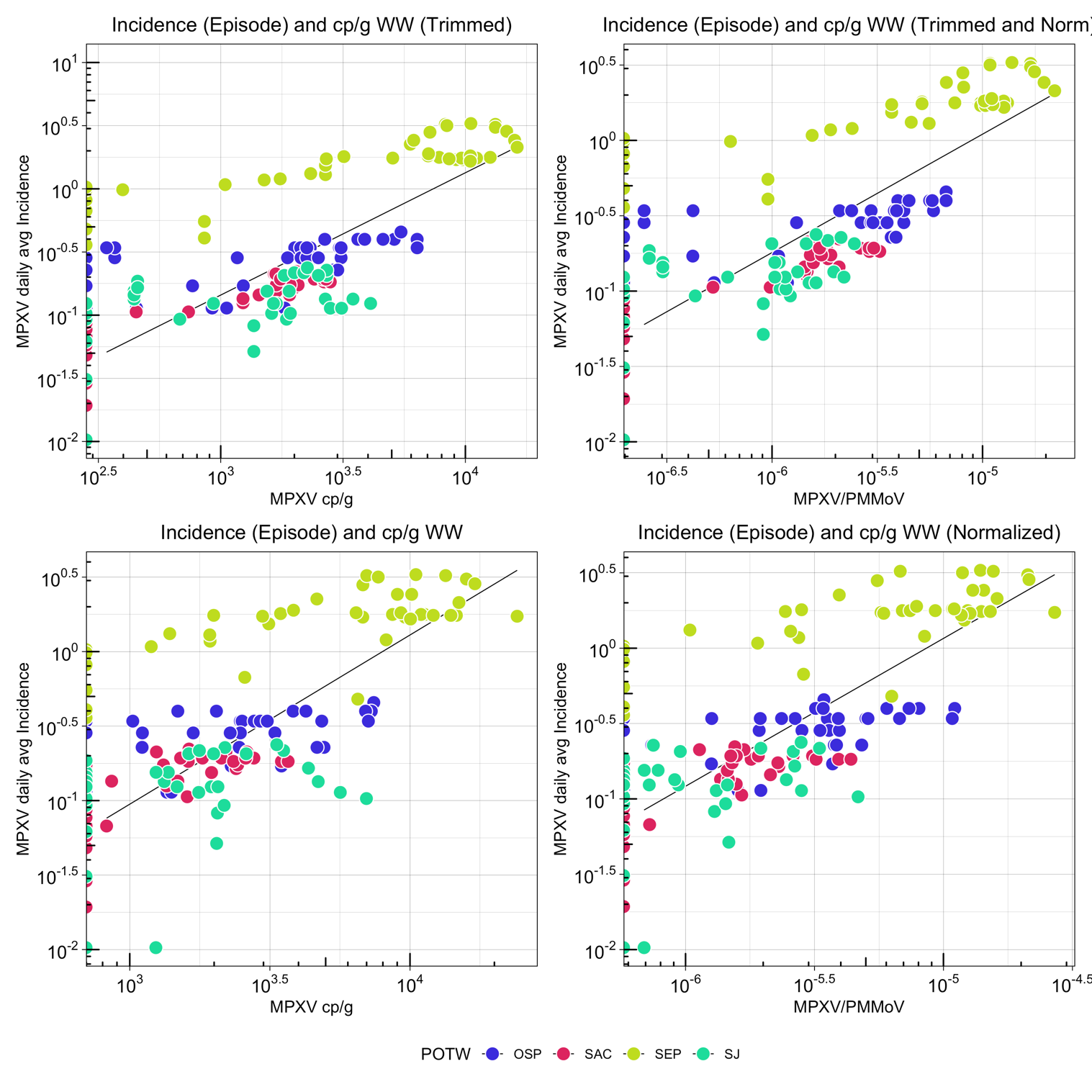

Fig S1. Association between cases (using episode date) and wastewater.

Monkeypox 7-day smoothed incidence rate (by episode date) plotted against MPXV data from wastewater(MPXV cp/g and MPXV normalized by PMMoV, and with and without 5-day trimmed smoothing). 8 points with a very low incidence rate < 10^-2^ (due to 0 zero reported cases in a large population) are not visualized. Of these, 7 were also non detect for MPXV DNA.

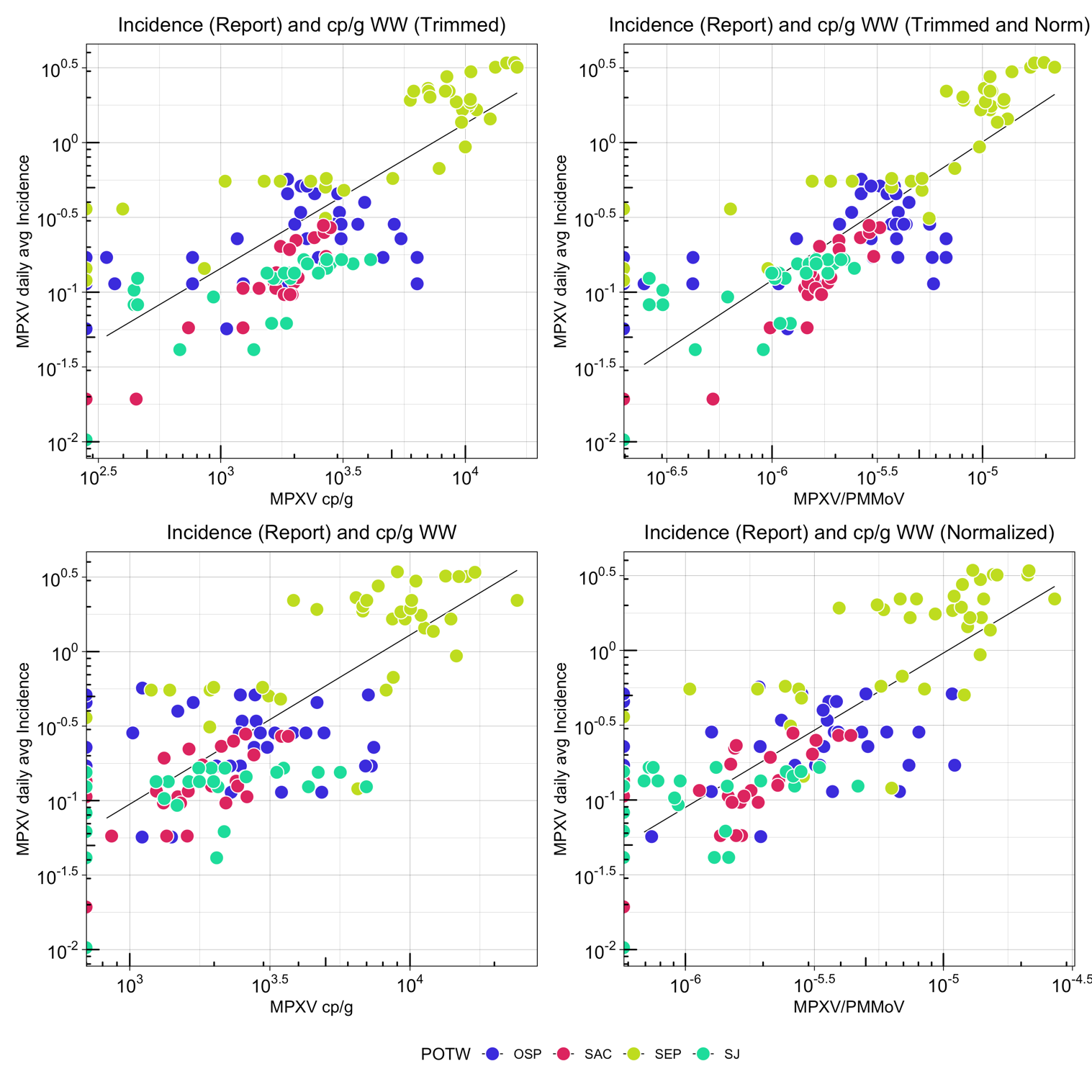

Fig S2. Association between cases (using report date) and wastewater.

Monkeypox 7-day smoothed incidence rate (by report date) plotted against MPXV data from wastewater(MPXV cp/g and MPXV normalized by PMMoV, and with and without 5-day trimmed smoothing). 8 points with very low incidence rate < 10^-2^ (due to 0 zero reported cases in a large population) are not visualized. 30 points with a very low incidence rate < 10^-2^ (due to 0 zero reported cases in a large population) are not visualized. Of these, 26 were also non-detect for MPXV DNA.

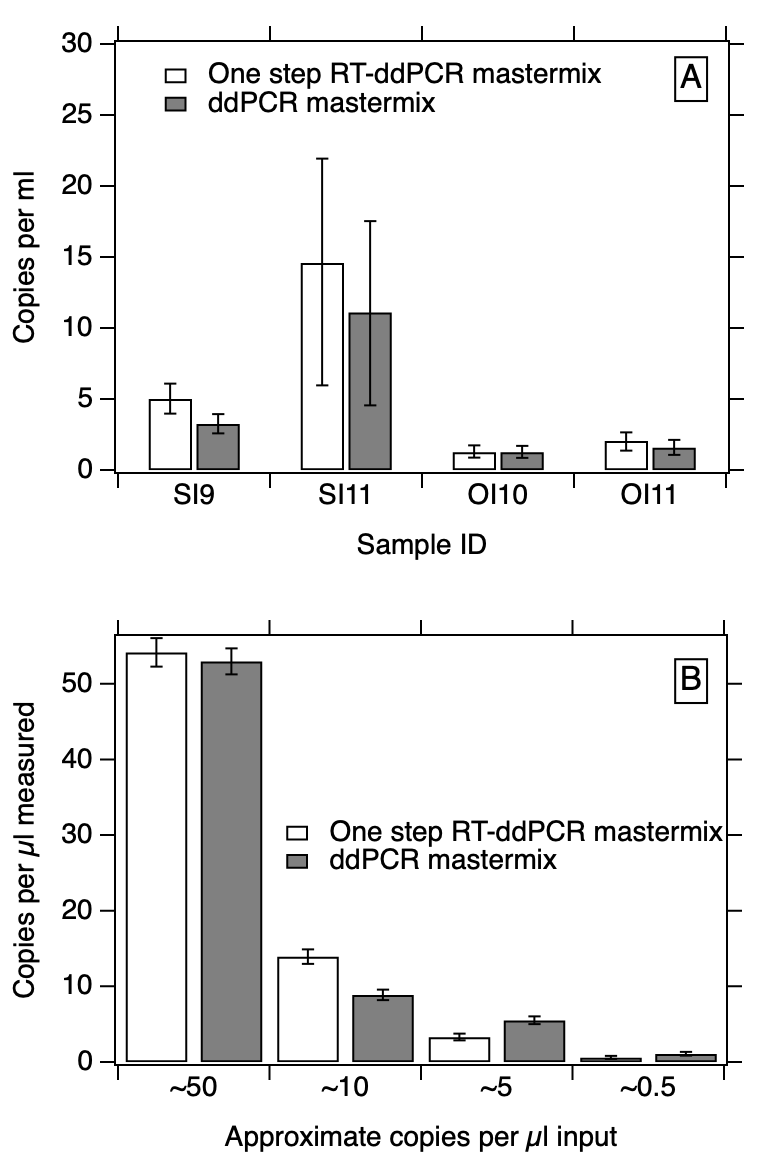

Figure S3. Panel A. Comparison between wastewater influent nucleic acid extracts (sample IDs provided) run using two different mastermixes using the same methods described in the paper for wastewater sample analysis. Panel B. Comparison between G2R_G standards (gene fragments) run using two different mastermixes. The approximate concentration of the standard input into the reaction is shown on the x-axis. Standards were run in three wells and the wells merged for analysis. The error bars represent the standard deviations as the total error reported from the instrument software and includes the Poisson error and the error among replicate wells.

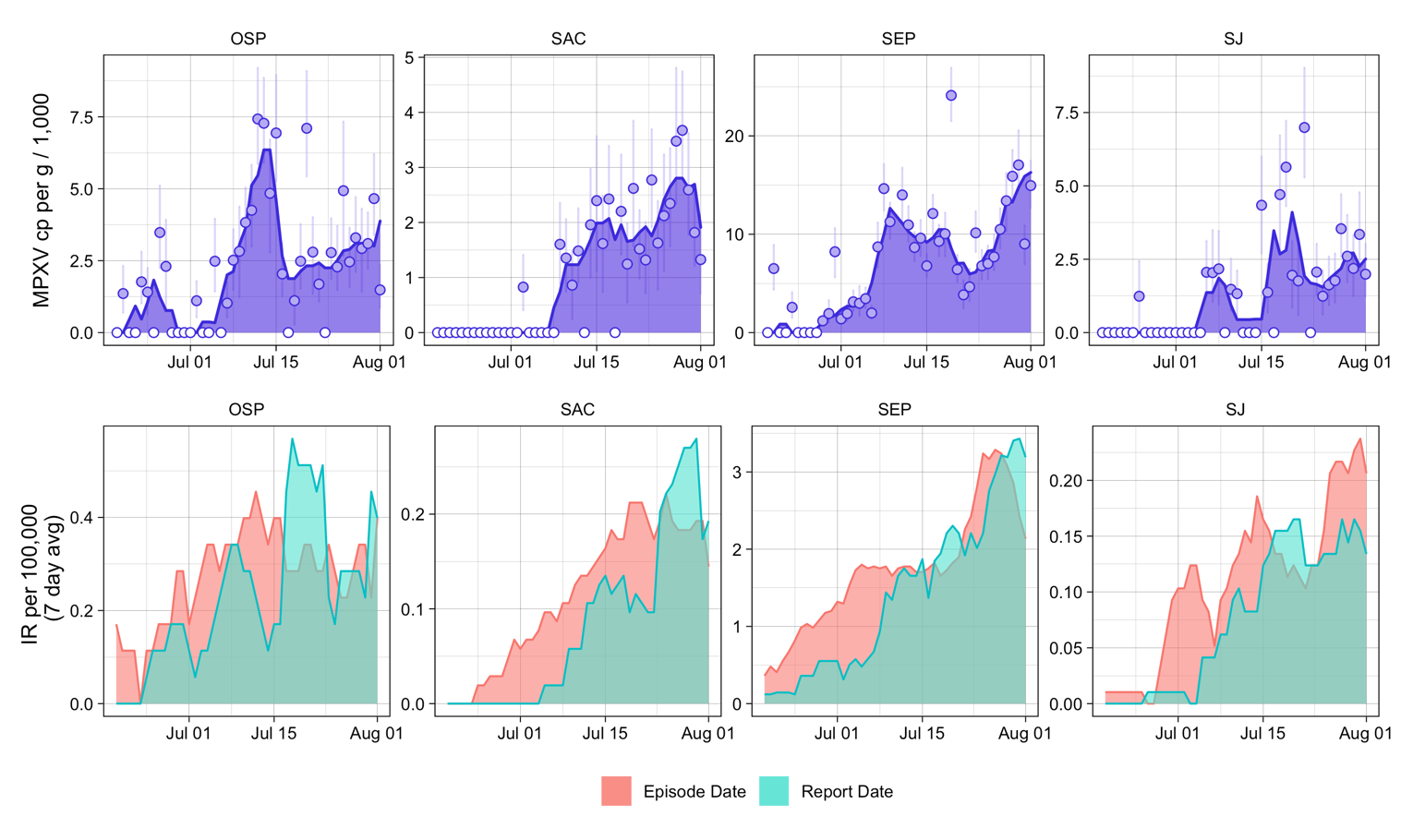

Fig S4. Wastewater monkeypox virus (MPXV) concentrations and incidence of monkeypox cases by sewershed. This figure contains the same data as Fig 2 in the main text, but with a free y axis to allow for closer examination in the patterns of cases and DNA in wastewater.

Top row: Time series of wastewater concentrations (concentration of MPXV DNA normalized by concentration of PMMoV RNA) at select publicly owned treatment works (POTWs) with >3 consecutive positive detections during the study time period: SEP (Southeast, San Francisco), OSP (Oceanside, San Francisco), Sac (Sacramento), and SJ (San Jose). The area under the curve represents the 5 day trimmed average of MPXV DNA cp/g over PMMoV cp/g in wastewater. Points represent daily values; open circles indicate non-detects. Error bars represent standard deviations and include Poisson error and variability among the 10 replicates (68% confidence intervals reported by the instrument software as “total error”). Bottom row: Daily incidence rate (IR) or monkeypox cases, averaged over 7-days, using episode date (red) and report date (green).
